# Genetic and maternal environmental contributions to estimated fetal weight at 20 weeks gestation compared with birthweight

**DOI:** 10.64898/2026.06.18.26355970

**Authors:** Rosie A. Purdy, Jingxue Feng, Zhenyu Luo, Robin N. Beaumont, Andrew T. Hattersley, Jianrong He, Xiu Qiu, Rachel M. Freathy, Alice E. Hughes

**Author notes:** <u>Corresponding author:</u> Rosie Purdy. Address: Department of Clinical and Biomedical Sciences, Faculty of Health and Life Sciences, University of Exeter, Exeter, UK.

## Abstract

**Introduction:** Birthweight reflects fetal growth from conception. The complications arising from the extremes of fetal growth are well known, but addressing these effectively depends on understanding how and when fetal growth is affected by various maternal and fetal factors. We aimed to compare associations of fetal genetic and maternal environmental factors with fetal weight mid-pregnancy and birthweight.

**Material and methods:** We studied 3110 mother-child pairs from three population-based birth cohorts (UK and China). Estimated fetal weight at ∼20 weeks gestation (EFW20) and birthweight were the outcomes of interest. Exposures were fetal genetic factors (fetal sex and fetal birthweight genetic score [BW GS]) and maternal factors (maternal BW GS, BMI, fasting plasma glucose [FPG], smoking, age, and parity). Associations were studied using multivariable linear regressions within cohorts and meta-analysed.

**Results:** All exposures were associated with both EFW20 and birthweight, apart from maternal FPG and smoking which were associated with birthweight only. Male fetal sex and a higher maternal BW GS were consistently associated with higher EFW20 and birthweight, whereas the fetal BW GS, maternal FPG, BMI and smoking showed more marked associations later in pregnancy. Maternal age and parity showed directionally opposite associations with EFW20 and birthweight.

**Conclusions:** Fetal genetic and maternal environmental factors vary in their effect on fetal growth in mid-pregnancy compared with late pregnancy. These findings contribute to our understanding of growth across pregnancy and may inform the timing of clinical monitoring of fetal growth and interventions targeting modifiable maternal factors.

**Key Message:** Drivers of fetal growth have been under-studied in their magnitudes and timings across gestation. We show that fetal genetic and maternal environmental factors vary in their associations with fetal growth at ∼20 weeks compared with later pregnancy.

## INTRODUCTION

Birthweight reflects the sum of fetal growth from conception to delivery. Individuals born with high or low birthweight are at higher risk (compared with those of average birthweight) of important perinatal complications^1^ and of adult-onset diseases, such as type 2 diabetes.^2^ A better understanding of fetal growth could therefore improve our understanding and management of multiple adverse health outcomes. Many factors are known to influence fetal growth and ultimately birthweight. These can be broadly divided into maternal physiological and environmental characteristics (eg glucose levels, BMI, smoking status), and fetal genetic factors, which may influence growth directly or via placental function/efficiency.^3^

The trajectory of fetal growth is not linear^4^ and is likely influenced by different factors at different times in gestation. For example, maternal glucose is a key regulator of fetal insulin-mediated growth and exerts its influence primarily in the third trimester,^5^ yet associations between fetal sex and fetal biometry are apparent from the first trimester.^6^ Fetal genetic factors, including fetal sex, generally reflect an individual’s inherent growth potential, which may have limited sensitivity to any subtle changes in placental efficiency as pregnancy progresses. In contrast, maternal environmental factors affect nutritional supply to the fetus, which may become more important as the placenta develops and drives fetal growth.^7^ However, for many factors that influence birthweight, their effects on fetal growth at different points in gestation are not well understood.

From a clinical perspective, knowing which factors are important at different stages of pregnancy would be useful to focus monitoring and treatment of modifiable factors at appropriate times, and for devising potential tools to predict high or low birthweight. In addition, such knowledge might inform future research into the mechanisms that influence fetal growth, helping us to understand why growth may deviate from normal.

In this study, we aimed to investigate the associations of different maternal characteristics and fetal genetic factors with estimated fetal weight at 20 weeks gestation, and to compare these with their associations with birthweight at term (≥37 weeks gestation) in three independent international cohorts. We hypothesized that fetal genetic factors would have consistent effects on growth across gestation, whereas maternal characteristics (genetic or environmental) that influence the intrauterine environment would have a greater effect on growth later in pregnancy.

## MATERIAL AND METHODS

We selected both fetal genetic factors and maternal genetic and physiological characteristics that influence the intrauterine environment (hereafter referred to as “maternal environmental factors”). Fetal genetic factors were fetal sex and fetal genetic score for birthweight (BW GS). A maternal BW GS was used to measure the impact of maternal genetic factors on fetal growth via the intrauterine environment, independently of those inherited by the child. Maternal environmental factors were body mass index (BMI), age, fasting plasma glucose (FPG) measured at ∼28 weeks gestation, smoking status and parity. The outcomes of interest were estimated fetal weight at 20 weeks gestation (EFW20, middle of pregnancy) and birthweight (BW, at term).

We constructed univariable and multivariable linear regression models with either EFW20 or birthweight as the outcome variable, and the genetic and environmental factors as predictor variables. We performed analyses within four separate samples of mother-child pairs (generated from three birth cohorts) and meta-analysed the results.

### a. Included cohorts/study samples

Born in Bradford (BiB) is a longitudinal multi-ethnic birth cohort study established in Bradford, UK, in 2007 to examine the impact of genetic, environmental, behavioural and social factors on child and maternal health and well-being in a deprived population.^8^ Data were originally collected from 12 453 women with 13 776 pregnancies, approximately half of whom were people of South Asian ethnicity (predominantly Pakistani), and half of non-South Asian ethnicity (predominantly White British).^8^ A total of 4081 pregnancies in unrelated mothers with genotype data were available for both mother and child.

The Exeter Family Study of Childhood Health (EFSOCH) is a prospective community-based study within central Exeter, UK, established in 2000 to investigate genetic and environmental influences on fetal growth and early development. Established inclusion criteria yielded a homogenous cohort of 1017 families (mother-father-child triads), comprised of non-diabetic mothers and fathers of self-reported UK Caucasian ethnicity.^9^ A total of 675 unrelated pregnancies had genotype data available for both mother and child.

The Born in Guangzhou Cohort Study (BIGCS) is a longitudinal birth cohort study based in the city of Guangzhou, southern China, launched in 2012 to explore how early-life traits impact developmental health.^10^ Data were originally collected from over 63 000 pregnancies from individuals of Chinese nationality. A total of 3851 pregnancies had genotype data available for both mother and child. Genotyping, imputation and kinship analyses are described in Supporting Information, Text 1.

### b. Selection of mother-child pairs for inclusion in analyses

We included singleton, term (37-42 weeks gestation) delivered pregnancies with complete maternal and fetal genotype data available. Women with pre-existing and pregnancy-induced hypertension, preeclampsia, and pre-existing diabetes (maternal FPG ≥7 mmol/L, and 2-hr glucose levels ≥11 mmol/L in BiB only) were excluded to create a clinically ‘healthy’ baseline cohort. Any pregnancies with missing variables included in our regression models were also excluded. Mother-offspring pairs in BiB were divided into separate analysis groups based on European and Pakistani genetic ancestry. A total of 3110 mother-child pairs remained after the filtering process, as outlined in Figure S1.

### c. Sample acquisition / measurements

Estimated fetal weight was calculated using Hadlock’s formula (see Supporting Information, Text 2) and adjusted for gestational age at ultrasound scan. Ultrasound measurements of head circumference, femur length and abdominal circumference were taken from the closest scan to 20 weeks gestation, between 18-22 weeks. This window was extended to include scans up to 25 weeks in BIGCS, due to 22-24 weeks being the common period for anomaly screening in China. Babies were weighed within 12 hours of delivery.^10^

Maternal FPG was measured during a standard oral glucose tolerance test at ∼28 weeks gestation in BiB and EFSOCH and 24-28 weeks in BIGCS. Maternal weight was measured at ∼28 weeks of gestation and used with measured (BiB and EFSOCH) or self-reported height (BIGCS) to calculate maternal BMI: weight (kg) / (height in m)^2^. Data was collected from participant questionnaires or medical records on maternal age, parity (number of previous pregnancies resulting in livebirths) and smoking status. Parity was coded as a binary primiparous/multiparous status. We coded to a binary smoker or nonsmoker status according to whether the mother smoked in pregnancy or not.

We calculated fetal and maternal genetic scores for birthweight (BW GSs) by summing birthweight-raising alleles of 216 SNPs in each individual and weighting by the beta values that were reported with robust evidence of genome-wide significance in the largest published GWAS meta-analysis of birthweight^11^ and available in all BiB, BIGCS, and EFSOCH study samples (see ST1). Further details for calculating these genetic scores are provided in the Supporting Information, Text 3.

### d. Statistical analyses

Analyses were performed in R (v4.3.2). Continuous variables (fetal BW GS, maternal BW GS, maternal BMI, maternal FPG, and maternal age) were converted to z-scores (standardized within each study sample to have a mean of 0 and SD of 1), and outlier entries (>+/-5 SD) were removed. Gestational-age-adjusted z-scores for birthweight and EFW20 were derived as standardized residuals from linear regression models of each measure on gestational age the measurement took place. These z-scores were used as outcomes in subsequent analyses, and gestational age was not included as a covariable. Multivariable linear regression models were performed within each study sample, with either gestational age-corrected EFW20 or birthweight z-score as the outcome of interest. These fetal and maternal models were constructed as follows:

Fetal models: EFW20/BW ∼ Fetal BW GS + fetal sex + first 5 PCs + genotyping chip/batch Maternal models: EFW20/BW ∼ Maternal BW GS + first 5 PCs + genotyping chip/batch + maternal BMI + maternal age + maternal FPG + parity status + smoking status

An extremely low number of Pakistani and Chinese mothers in this study were reported smokers (n=29 and 13, respectively), so we excluded the smoking variable from Pakistani and Chinese maternal models. A random-effects meta-analysis was performed using R package “metafor”^12^ to estimate overall associations of the predictor variables with each outcome across all four samples (total n=3110). Statistics to assess heterogeneity included I^2^ and Cochrane’s Q-statistic. Separate regression models by each predictor of interest were also performed. Analysis of BIGCS mother-child pairs was performed separately by the BIGCS team, and summary results were shared for the meta-analysis.

### e. Sensitivity analyses

We conducted sensitivity analyses to assess the influence on our results of gestational diabetes or the inclusion of ultrasound scan data later than 24 weeks. Full details of these analyses are in Supporting Information, Text 4.

## RESULTS

### Characteristics of included mother-offspring pairs

Profiles of each cohort’s mother-offspring pairs are shown in Table 1.

**Table 1.**
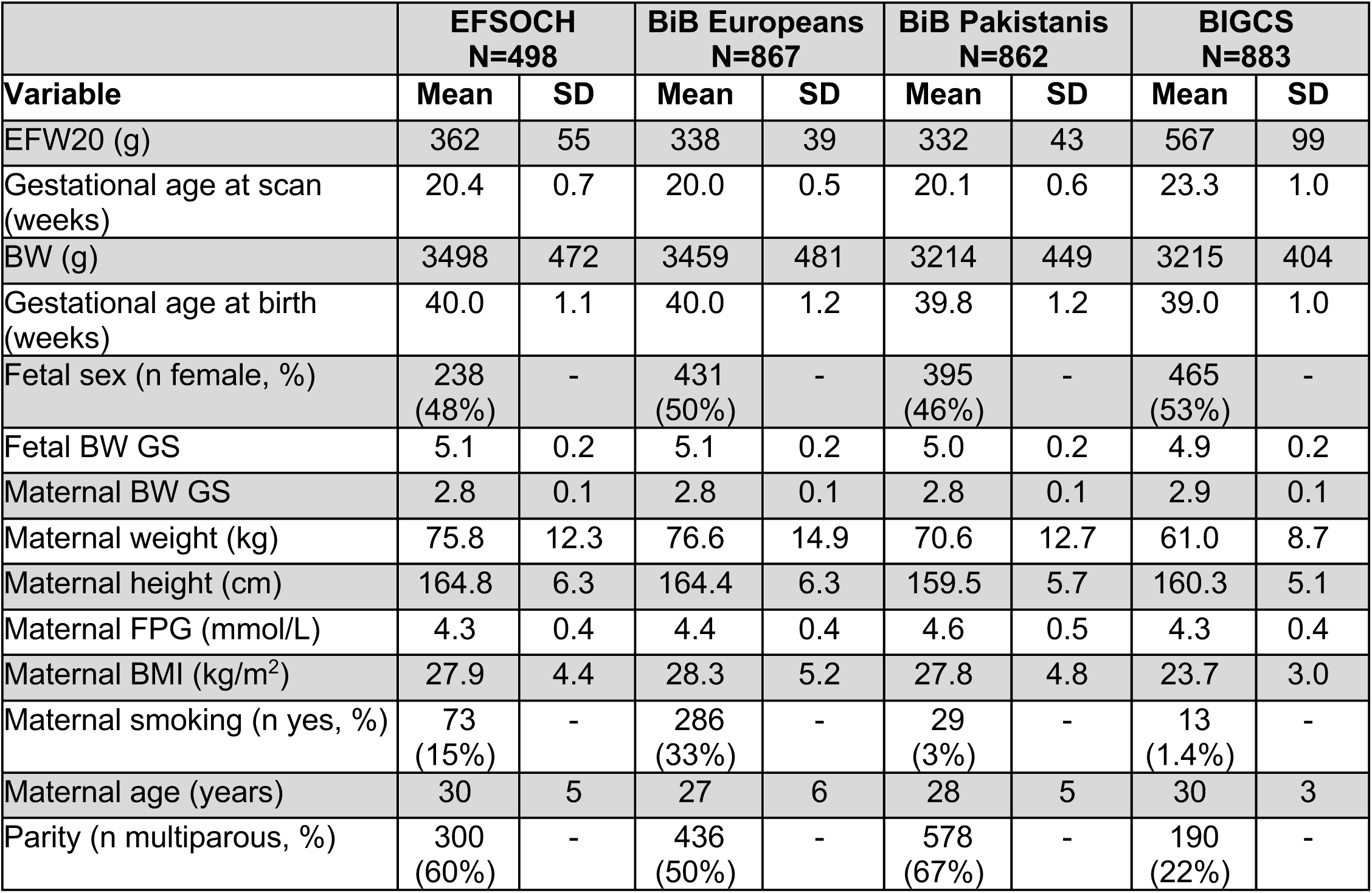
Characteristics of the 3110 mother-offspring pairs in the three study samples. EFW20: estimated fetal weight at ∼20 weeks gestation; BW: birthweight; BW GS: birthweight genetic score; FPG: fasting plasma glucose; BMI: body mass index.

|  | **EFSOCH**<br>**N=498** |  | **BiB Europeans**<br>**N=867** |  | **BiB Pakistanis**<br>**N=862** |  | **BIGCS**<br>**N=883** |  |
| --- | --- | --- | --- | --- | --- | --- | --- | --- |
| **Variable** | **Mean** | **SD** | **Mean** | **SD** | **Mean** | **SD** | **Mean** | **SD** |
| EFW20 (g) | 362 | 55 | 338 | 39 | 332 | 43 | 567 | 99 |
| Gestational age at scan (weeks) | 20.4 | 0.7 | 20.0 | 0.5 | 20.1 | 0.6 | 23.3 | 1.0 |
| BW (g) | 3498 | 472 | 3459 | 481 | 3214 | 449 | 3215 | 404 |
| Gestational age at birth (weeks) | 40.0 | 1.1 | 40.0 | 1.2 | 39.8 | 1.2 | 39.0 | 1.0 |
| Fetal sex (n female, %) | 238<br>(48%) | - | 431<br>(50%) | - | 395<br>(46%) | - | 465<br>(53%) | - |
| Fetal BW GS | 5.1 | 0.2 | 5.1 | 0.2 | 5.0 | 0.2 | 4.9 | 0.2 |
| Maternal BW GS | 2.8 | 0.1 | 2.8 | 0.1 | 2.8 | 0.1 | 2.9 | 0.1 |
| Maternal weight (kg) | 75.8 | 12.3 | 76.6 | 14.9 | 70.6 | 12.7 | 61.0 | 8.7 |
| Maternal height (cm) | 164.8 | 6.3 | 164.4 | 6.3 | 159.5 | 5.7 | 160.3 | 5.1 |
| Maternal FPG (mmol/L) | 4.3 | 0.4 | 4.4 | 0.4 | 4.6 | 0.5 | 4.3 | 0.4 |
| Maternal BMI (kg/m<sup>2</sup>) | 27.9 | 4.4 | 28.3 | 5.2 | 27.8 | 4.8 | 23.7 | 3.0 |
| Maternal smoking (n yes, %) | 73<br>(15%) | - | 286<br>(33%) | - | 29<br>(3%) | - | 13<br>(1.4%) | - |
| Maternal age (years) | 30 | 5 | 27 | 6 | 28 | 5 | 30 | 3 |
| Parity (n multiparous, %) | 300<br>(60%) | - | 436<br>(50%) | - | 578<br>(67%) | - | 190<br>(22%) | - |

### Association between birthweight and fetal weight at 20 weeks

Estimated fetal weight at 20 weeks was associated with birthweight (meta-analysis effect estimate: 0.33 SD higher EFW20 per 1 SD higher birthweight, 95% confidence interval [0.27,0.39 SD], *p*-value = 4.16 x10^-25^, see ST2 & Figure S2).

### Associations of fetal genetic factors with fetal growth

#### Fetal sex

Females were lighter on average at 20 weeks gestation (−0.27 [-0.35,-0.19] SD, *p*-value = 2.79 x10^-11^) and when born (−0.32 [-0.39,-0.26] SD, *p* = 1.08 x10^-20^) than males (see Fig. 1 & ST3). These results correspond to a difference of approximately −32 g and −150 g for EFW20 and BW, respectively. Overall little heterogeneity was observed in the pooled fetal sex effect estimates for EFW at 20 weeks (I^2^ = 23%) and for birthweight (I^2^ = 0%; ST4).

**Figure 1.**
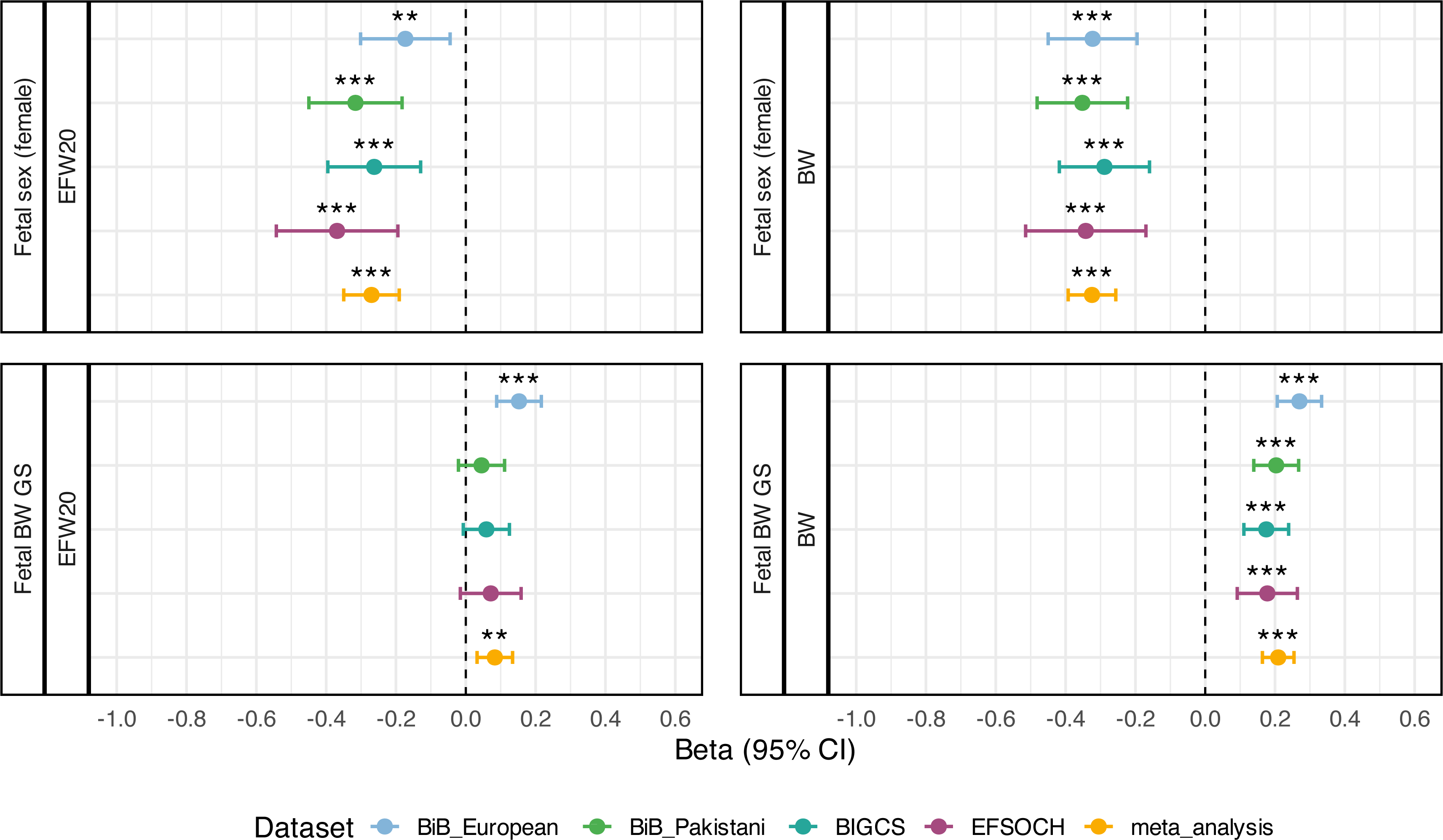
Associations between fetal genetic factors and fetal growth outcomes at 20 weeks gestation and at term. The beta value on the x-axis represents the standard deviation unit change in estimated fetal weight (EFW20) or birthweight (BW) per 1 unit change in the standardized or binary exposure variable. * = *p* <0.05, ** = *p* <0.01 and *** = *p* <0.001.

#### Fetal BW GS

In the meta-analysis, a higher fetal BW GS was associated with a higher EFW20 (0.08, [0.03,0.13] SD, *p* = 1.35 x10^-3^) and a higher birthweight (0.21 [0.16,0.25] SD, *p* = 1.43 x10^-19^, Fig. 1 & ST3). These effect estimates were equivalent to 10 g and 98 g for EFW20 and BW, respectively, per 1 SD increase in fetal BW GS. Both meta-analysed results were directionally consistent but the association with BW had a greater magnitude. Moderate heterogeneity was observed in the pooled fetal BW GS effect estimates for EFW20 (I^2^=53%), and BW (I^2^=42%; ST4).

### Associations of maternal environmental factors with fetal growth

Of the six maternal environmental factors we investigated, maternal BW GS showed consistent effects on fetal growth at mid-pregnancy and term, maternal FPG, BMI and smoking showed more marked effects later in pregnancy, and maternal age and parity showed directionally opposing effects on growth when comparing their effects on EFW20 and BW (Fig. 2).

**Figure 2.**
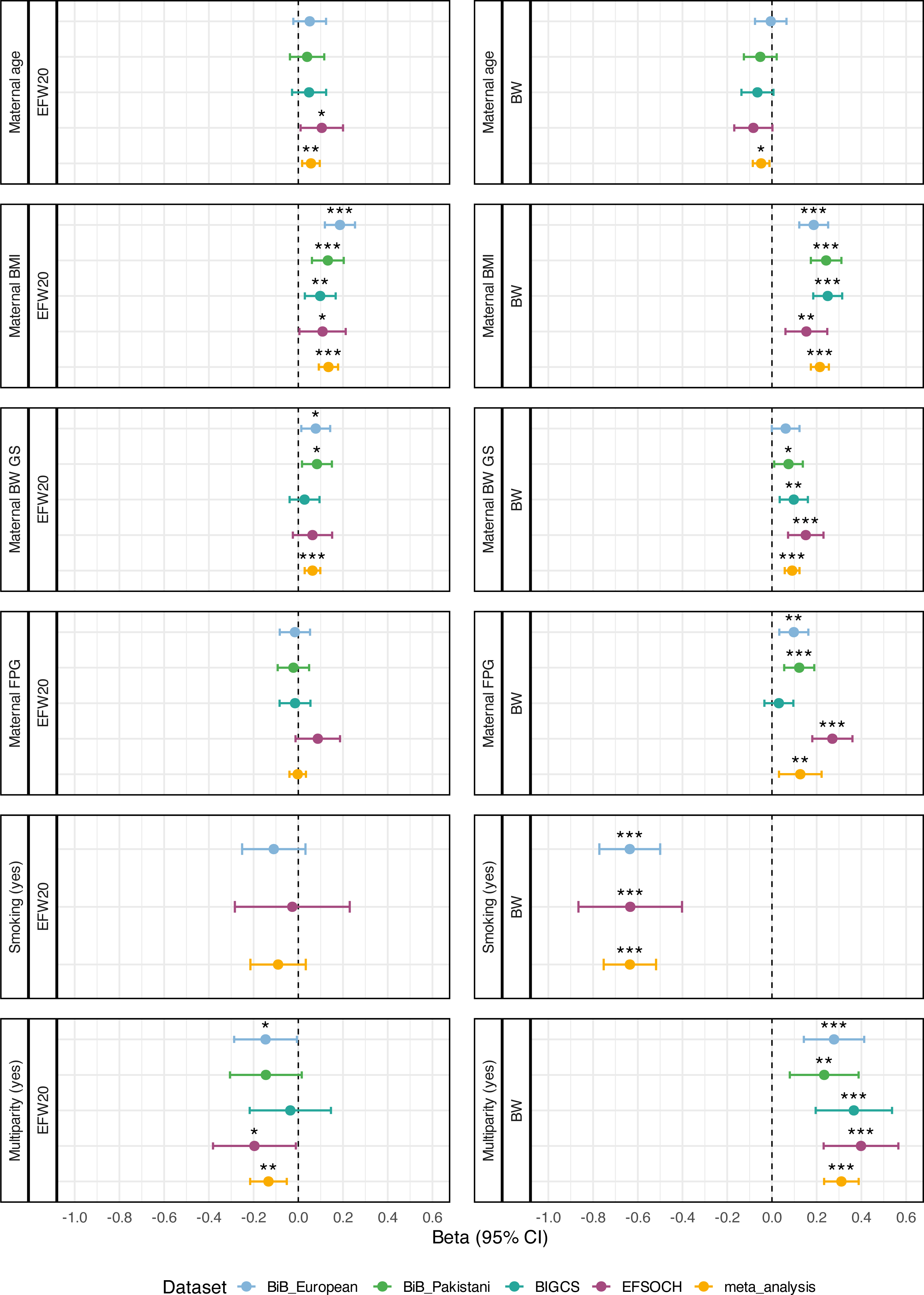
Associations between maternal factors and fetal growth outcomes at 20 weeks gestation and at term. The beta value on the x-axis represents the standard deviation unit change in estimated fetal weight at 20 weeks gestation (EFW20) or birthweight (BW) per 1 unit change in the standardized or binary exposure variable. * = *p* <0.05, ** = *p* <0.01 and *** = *p* <0.001. Maternal smoking was not included in the BiB Pakistani and BIGCS maternal models due to low sample size of smokers.

#### Maternal BW GS

Maternal genetics, acting through the intrauterine environment, had a consistent effect on fetal growth at both time points: a higher maternal BW GS had a positive association with both EFW20 (0.06, [0.03,0.10] SD, *p* = 3.64 x10^-4^) and birthweight (0.09 [0.06,0.12] SD, *p* = 7.88 x10^-8^, see Fig. 2 & ST3), corresponding to a 7 g and 42 g higher weight per 1 SD higher maternal BW GS, respectively. Pooled effect estimates showed homogeneity at both 20 weeks (I^2^ = 0%) and at term (I^2^=13%; ST4).

#### Maternal BMI

A higher maternal BMI was associated with a higher EFW20 (0.13 [0.09,0.18] SD, *p* = 8.49 x10^-10^) and higher birthweight to a greater degree (0.21 [0.17,0.25] SD, *p* = 2.03 x10^-25^, Fig. 2 & ST3). This is equivalent to a 17 g and 98 g higher weight per 1 SD higher maternal BMI, respectively. There was little evidence of heterogeneity across cohorts at both points in pregnancy (EFW20 I^2^=24%; BW I^2^=23%; ST4).

#### Maternal glucose

There was no association between higher maternal FPG level and fetal weight at 20 weeks (−0.003 [-0.04,0.03] SD, *p* = 0.891), with all analysis groups’ effect estimates crossing the null. However, there was strong evidence of an association between maternal FPG and higher birthweight (0.13 [0.03,0.22] SD, *p* = 9.07 x10^-3^, Fig. 2 & ST3), an increase of approximately 61 g per 1 SD increase in maternal FPG. Heterogeneity was observed in the pooled FPG effect estimates for birthweight (I^2^=86%; Q *p* = 3.84 x10^-4^; ST4). The FPG–BW association was stronger in EFSOCH than in BiB Europeans (difference=80 g per 1 SD increase in FPG, SE=27g, z=2.99, *p* = 0.003). Also, the FPG–BW association in the BIGCS group crossed the null in the multivariable model (0.03 [-0.03,0.10] SD, *p* = 0.349; ST3), but there was a statistically significant positive association in the univariable model (0.104 [0.04,0.17] SD, *p* = 1.99 x10^-3^, see ST5).

After applying more stringent glucose criteria to our analyses to reduce potential bias from post-diagnosis GDM treatment, 148 mother-child pairs were excluded, totalling 2962 samples left for sensitivity analysis. These meta-analysed results were highly consistent with the main analysis, with only slight attenuations in associations between maternal environmental factors and EFW20 and birthweight (see ST6,7 & Fig S3). The additional sensitivity analysis, which restricted the ultrasound window in the BIGCS cohort to 18-24 weeks (n=704 mother-child pairs, n=179 from main analysis excluded) gave similar results as the main analysis in their direction and magnitudes of associations with EFW20 (see ST8).

#### Maternal smoking

We found no evidence of an association between maternal smoking during pregnancy and with EFW20 in participants of European ancestry (−0.09 [-0.21,0.03] SD, *p* = 0.153, I^2^=0). However, smoking was strongly associated with a lower birthweight (−0.64 [-0.75,-0.52] SD, *p* = 2.07 x10^-26^, I^2^=0%; ST3 and ST4), the equivalent of babies born to mothers who smoked during pregnancy being 300 g lighter than nonsmokers of these groups.

#### Multiparity

Meta-analysed results showed that multiparity was associated with a lower fetal weight at 20 weeks (−0.13 [-0.22,-0.05] SD, *p* = 1.40 x10^-3^, I^2^=0%). In contrast, being multiparous was associated with having a baby with a higher birthweight (0.31 [0.23,0.39] SD, *p* = 3.01 x10^-15^, I^2^=0%; ST3 and ST4). This corresponds to fetuses from multiparous mothers being 16 g lighter than fetuses from first-time mothers at 20 weeks, but at term these babies were born 145 g heavier than babies from first-time mothers.

#### Age

A higher maternal age was associated with a small increase in fetal weight at 20 weeks (0.06 [0.02,0.10] SD, *p* = 4.88 x10^-3^, I^2^=0), but a small decrease in birthweight (−0.05 [-0.09,-0.01] SD, *p* = 1.09 x10^-2^, I^2^=0; ST3 and ST4). This equates to a 7 g higher EFW20, and a 23 g lower BW per 1 SD higher maternal age.

## DISCUSSION

This study investigated fetal genetic and maternal environmental contributions to earlier and later fetal growth and found differing associations. The results were consistent across three international cohorts of diverse genetic ancestry and provide interesting insights into the factors that govern fetal growth in humans.

It is well known that females are lighter than males at birth. We observed them to have a lighter estimated fetal weight than males at 20 weeks gestation, consistent with results from other studies,^13, 14^ with sexual size dimorphism being observed as early as 8-12 weeks.^15^ Sex-specific differences in fetal growth patterns may be attributed to distinct variations in placentation,^16^ endocrine signalling^17^ and genetics.^18^

Regarding the fetal BW GS, the influence of birthweight-raising-alleles on placental function is likely to contribute to the difference in the magnitude of fetal growth effects at 20 weeks compared to at term. A proportion of the birthweight genetic scores will drive placental weight (PW; a proxy for placental growth); of the 33 signals (41 loci) identified in a PW GWAS meta-analysis,^19^ seven of these variants are present in the set of 216 birthweight-raising-alleles used to construct the GSs for this study, and a further 23 variants from the PW GWAS^19^ were located in close proximity to birthweight variants also used for this study. There is a strong positive correlation between birthweight and PW,^19^ which captures non-genome-wide significant SNPs. Fetal insulin regulates PW,^19^ and evidence supports several birthweight-associated SNPs impacting insulin-mediated growth. These genetic factors should have a more marked effect on growth later in pregnancy, given that fetal insulin-mediated growth occurs primarily in the third trimester.^5^ Contrary to our study, a longitudinal analysis by Vermeulen et al.^20^ found no associations between a fetal BW GS and fetal growth measures, including EFW at 20 weeks, but they did detect associations with all measures at 30 weeks. However, this could be due to their analyses using an older GWAS, with fewer genome-wide significant SNPs (n=60) used in their GS compared to this study, thus limiting power to detect associations as early as 20 weeks. The 216 variants that made up our fetal genetic score were identified in a GWAS of birthweight, so the smaller association with earlier fetal growth is perhaps not surprising. This fetal BW GS–EFW20 association we observed is consistent with the fetal genetic score reflecting a mixture of constitutional and intrauterine-specific growth.^5, 7, 19^

The maternal BW GS was weighted by maternal effects, which are independent of fetal effects. So, although genetic, it reflects the effects of maternal genes whose effects on fetal growth are exerted via the intrauterine environment. The consistent association that we observed between the maternal BW GS and weight at mid-pregnancy and term is, to our knowledge, a novel finding. Although some of the maternal genetic variants are known to act via maternal fasting glucose,^19^ further work is needed to understand the other pathways affected. It is possible that the maternal BW GS is capturing some maternal genetic factors that influence early placental function; abnormal development of uteroplacental circulation forms the basis of fetal growth restriction etiopathology.^21^ Interestingly, a birthweight-lowering maternal GS was previously associated with lower EFW from 20 to 40 weeks.^22^

Maternal glucose crosses the placenta and stimulates fetal insulin secretion, which in turn accelerates fetal growth and enhances offspring adiposity at birth.^23^ This link between higher maternal glucose and offspring birthweight is well known and corroborated by the current study.

Other studies have shown changes in fetal growth in pregnancies later affected by GDM before 20 weeks,^24, 25^ however, the absence of effect of maternal FPG on fetal growth in mid-pregnancy in this study is likely explained by insulin-mediated growth being more important in the third trimester.^5^ Our sensitivity analysis where we excluded women who may have been treated for GDM and therefore had their glucose levels lowered also showed directionally consistent results to our main findings. The reason for the stronger FPG–BW association in EFSOCH compared to BiB Europeans is unclear but could be a result of socioeconomic differences between Bradford and Exeter which have not been accounted for in this study. Maternal FPG was not associated with birthweight in the multivariable BIGCS model, but it was in a univariable model, suggesting that the effect of glucose in that cohort was attenuated or obscured by other maternal factors. Overall, our results suggest that higher FPG levels are not driving increased fetal size in mid-pregnancy. Therefore, if a fetus appears large for its gestational age at 20 weeks, its growth is unlikely to be explained by maternal hyperglycaemia at that stage. Although it has been shown that treating maternal hyperglycemia earlier than 24 weeks limits excess fetal growth,^26^ our results do not support performing oral glucose tolerance tests at 20 weeks to detect hyperglycemia in women with large fetuses.

Elevated maternal BMI was associated with increased fetal weight at 20 weeks and at term. Raised maternal BMI is linked to higher maternal glucose.^23^ Maternal BMI had a greater impact on birthweight than fetal weight at 20 weeks, which could partly reflect BMI capturing maternal glucose that is not completely accounted for by FPG levels, or other factors not covered by this study, such as maternal lipid levels. Maternal lipids (such as triglycerides and cholesterols) are also important for fetal growth^27^ by providing energy for maternal metabolism and fetal development.^28^

We observed maternal tobacco-smoking was strongly associated with birthweight, which corroborates the well-established link between smoking during pregnancy and fetal growth restriction, resulting in low birthweight.^29^ In contrast, we found no significant association with overall fetal weight at around 20 weeks. A study by Jaddoe et al.^30^ revealed marked differences between biometrics of fetuses whose mothers continued to smoke after discovering they were pregnant compared to nonsmokers, and these differences widened with increased gestational age: maternal smoking was associated with smaller fetal femur length from 18-24 weeks onwards, and smaller head and abdominal circumference from ≥25 weeks and onwards.^30^ Such changes, if present in our cohorts, did not contribute to any detectable associations between maternal smoking and EFW20.

Multiparous women tend to have heavier offspring from the second pregnancy onwards compared to their firstborn.^31^ We confirmed a consistent positive association between multiparity and birthweight in our study groups. This is thought to occur due to structural and vascular adaptations to the uterus during the first pregnancy which do not fully reverse postpartum, consequently improving fetal nutrient delivery in subsequent pregnancies.^31^ Conversely, our results show evidence that being multiparous is associated with a lower fetal weight at 20 weeks. To our knowledge, there is no published evidence to date to suggest this negative association; in fact, in an analysis of 9031 mother-child pairs, Gaillard et al.^32^ observed heavier EFW20 on average in multipara compared to primipara. However, in that study, the difference was more pronounced for women with two or more previous pregnancies; fetuses of women with one previous pregnancy had smaller head circumference, on average, compared with those from those of nulliparous women. The clinical implications of parity for fetal growth would benefit from further study.

We observed an opposing association between older maternal age and higher EFW20 and lower birthweight.^33^ Advanced maternal age (defined as ≥35 years) has been linked to both increased risks of low birthweight^34^ and macrosomia.^35^ Other studies have also linked small increases in EFW20 with older maternal age,^33^ contrary to our results. The effect of maternal age on both EFW20 and term birthweight were very small, so unlikely to be clinically important, but it raises the possibility of a different trajectory of growth in pregnancies in older vs younger women.

This study used a high number of mother-child pairs from three international cohorts, with consistent results across analysis groups. The pooled cohorts represent an ethnically heterogenous sample, with a 56% non-European majority. Controlling for GDM post-diagnosis treatment still gives consistent results, which robustly suggests that this has not altered our conclusions about fetal growth. We did not have earlier scan measurements to make comparisons with the first trimester, but to directly compare those with term birthweight would be difficult, as fetal weight is not typically estimated until the late second trimester. Serial measurements of growth from the second trimester may have been helpful to study where shifts in the regulation of growth occur. Nevertheless, there may be lower sensitivity in detecting differences in fetal sizes at 20 weeks gestation compared to at term, which may have limited power to detect effects from our predictors of interest earlier on in pregnancy.

The window of ultrasound scans accepted in the BIGCS cohort (18-25 weeks) is wider than those from the UK, due to significantly fewer ultrasounds being performed at 18-22 weeks (n=81). From 24 weeks onwards, growth is increasingly driven by nutrition, and hence by the placenta,^36^ but we opted to include up to 25 weeks in BIGCS for a sufficient sample size. Despite these differences between BIGCS and the other cohorts, their associations remained mostly consistent. Furthermore, the sensitivity analysis restricting this window to 18-24 weeks in BIGCS yielded consistent results to the main analysis (ST8).

## CONCLUSION

In this study we have shown that fetal genetic and maternal environmental factors vary in their effect on fetal growth in mid-pregnancy compared to late pregnancy. Further research is required to clarify the mechanisms underlying growth across gestation and to subsequently manage pregnancies with abnormally projected fetal growth. Our findings provide further insight into the contributions of fetal genetics and intrauterine environment factors to fetal growth.

## Ethics Statement

Ethical approval for EFSOCH was given by the North and East Devon (UK) Local Research Ethics Committee (approval number 1104), and informed consent was obtained from the parents of the newborns. Ethics approval was obtained for the main platform study and all the individual sub-studies from the Bradford Research Ethics Committee. BIGCS was approved by Guangzhou Women and Children’s Medical Center Ethic Committee, and all participants provided written consent.

## Author contributions (CReDIT taxonomy)

**Rosie Purdy:** Data curation, formal analysis, investigation, visualisation, writing – original draft.

**Jingxue Feng:** Data curation, formal analysis, investigation, writing – review and editing.

**Zhenyu Luo:** Data curation, formal analysis, investigation, writing – review and editing.

**Robin Beaumont:** Methodology, software, supervision.

**Andrew Hattersley:** Conceptualization, supervision, writing – review and editing.

**Jianrong He:** Funding acquisition, supervision, writing – review and editing.

**Xiu Qiu:** Funding acquisition, supervision, writing – review and editing.

**Rachel Freathy:** Methodology, investigation, supervision, funding acquisition, writing – original draft, writing – review and editing.

**Alice Hughes:** Conceptualization, data curation, methodology, investigation, supervision, writing – original draft, writing – review and editing.

## Data availability

Summary statistics from EFSOCH are available on request. Researchers interested in accessing the data are expected to send a reasonable request by sending an email to the Exeter Clinical Research Facility at. Scientists are encouraged and able to use BiB data. Data requests are made to the BiB executive using the form available from the study website https://borninbradford.nhs.uk/our-data/how-to-access-data/. Guidance for researchers and collaborators, the data dictionary and data summary are all available via the website. All requests are carefully considered and accepted where possible. Researchers who are interested in collaborating with the BIGCS group are welcome to contact us via email at.

## Acknowledgements

We are grateful to all of the families who contributed to EFSOCH and the team who collected and curated the data. BiB is only possible because of the enthusiasm and commitment of the children and parents in BiB. We are grateful to all the participants, health professionals, schools and researchers who have made Born in Bradford happen. We would like to thank all the families who have participated in BIGCS, as well as the staff members for their dedication to data collection and management.

## Abbreviations

BiB: Born in Bradford
BIGCS: Born in Guangzhou Cohort Study
BMI: body mass index
BW GS: birthweight genetic score
EFSOCH: Exeter Family Study of Childhood Health
EFW20: estimated fetal weight at ∼20 weeks gestation
FPG: fasting plasma glucose
GDM: gestational diabetes mellitus

## Figure and Table Legends (Supporting Information)

**Table ST1. List of SNPs used in generating birthweight genetic scores (n=216).** EAF = Effect Allele Frequency; EGG-frq = Frequency of the effect allele in the EGG/UKB data; IS-frq = Frequency of effect allele in the Icelandic data.

**Table ST2. Association between estimated fetal weight and birthweight.** Beta value represents standard deviation unit change in EFW20 per 1 unit change in birthweight. SE: standard error. CI = 95% confidence interval. Meta-analysis: Random effects model (k=4).

**Table ST3. Multivariable model results, showing effects of multiple predictor variables on EFW20/BW, per dataset.** Beta value represents standard deviation unit change in EFW20 or BW per 1 unit change in the standardized or binary exposure variable. SE: standard error. CI = 95% confidence interval. EFSOCH n=498; BiB European n=867; BiB Pakistani n=862; BIGCS n=883.

**Table ST4. Meta-analysis results from metafor. Generated from multivariable models of main analyses.** Beta value represents standard deviation unit change in EFW20 or BW per 1 unit change in the standardized or binary exposure variable. SE: standard error. CI = 95% confidence interval.

**Table ST5. Univariable model results, showing effects of single predictor variables on EFW20/BW, per dataset.**

**Table ST6. ST6a. Sensitivity multivariable model results**, showing effects of multiple predictor variables on EFW20/BW with mothers with fasting glucose values ≥ 5.6 mmol/L and 2-hour glucose values ≥7.8 mmol/L (diagnostic of GDM), per dataset. **ST6b. Sensitivity univariable model results**, showing effects of predictor variables on EFW20/BW, with mothers with fasting glucose values ≥ 5.6 mmol/L and 2-hour glucose values ≥ 7.8 mmol/L (diagnostic of GDM), per dataset.

**Table ST7. Meta-analysis of glucose sensitivity results.** Beta value represents standard deviation unit change in EFW20 or BW per 1 unit change in the standardized or binary exposure variable. SE: standard error. CI = 95% confidence interval.

**Table ST8. Sensitivity multivariable model results, showing effects of multiple predictor variables on EFW20 in BIGCS with ultrasound scans restricted to 18-24 weeks gestation.** BIGCS n=704 after restricting ultrasound scan to 18-24 weeks.

**Figure S1.**
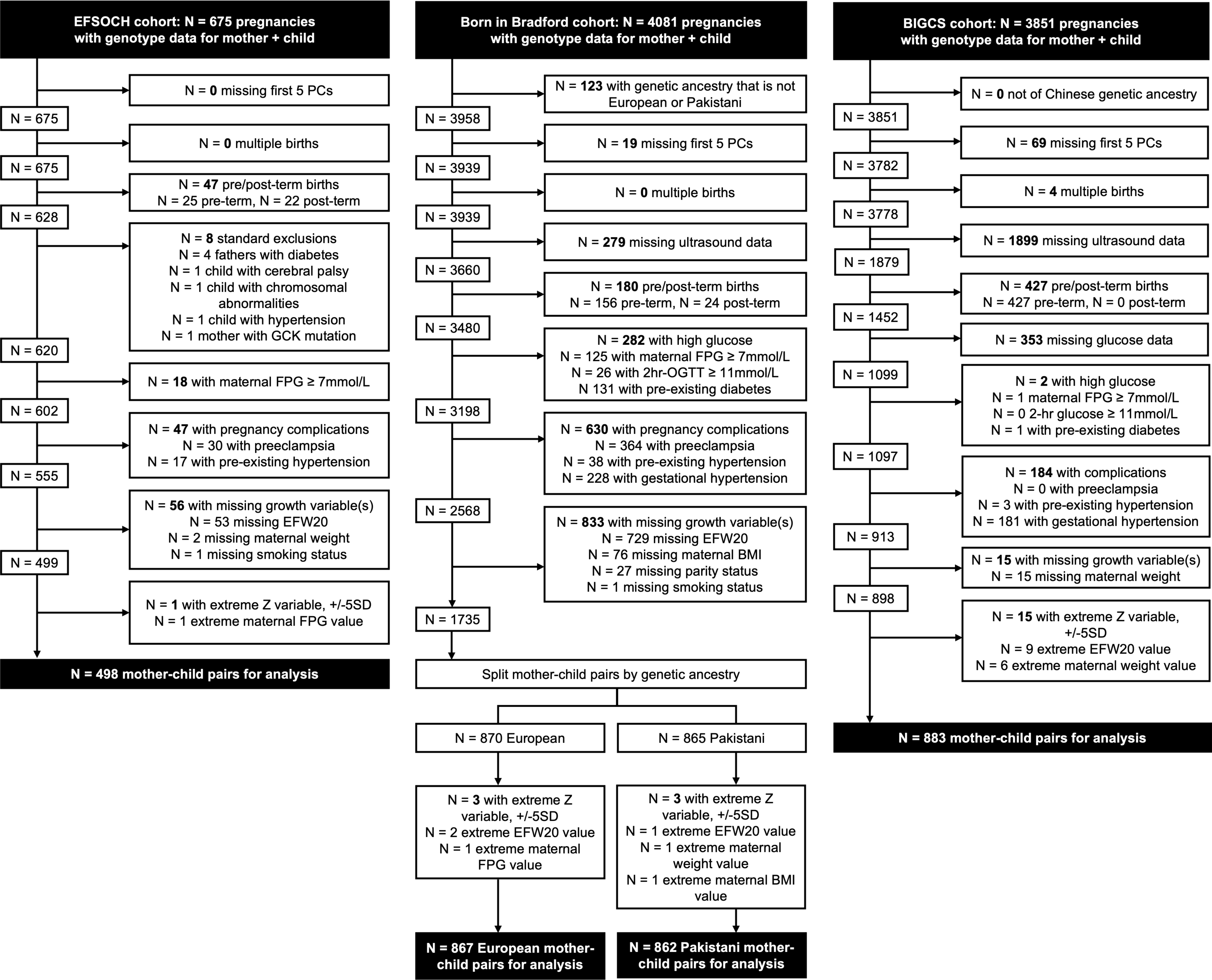
Flowcharts illustrating how the data was filtered and prepared for analysis. EFSOCH (left), BiB (middle) and BIGCS (right). No multiple births were reported when filtering the BiB dataset due to using data from unrelated families, of which only first births were selected.

**Figure S2.**
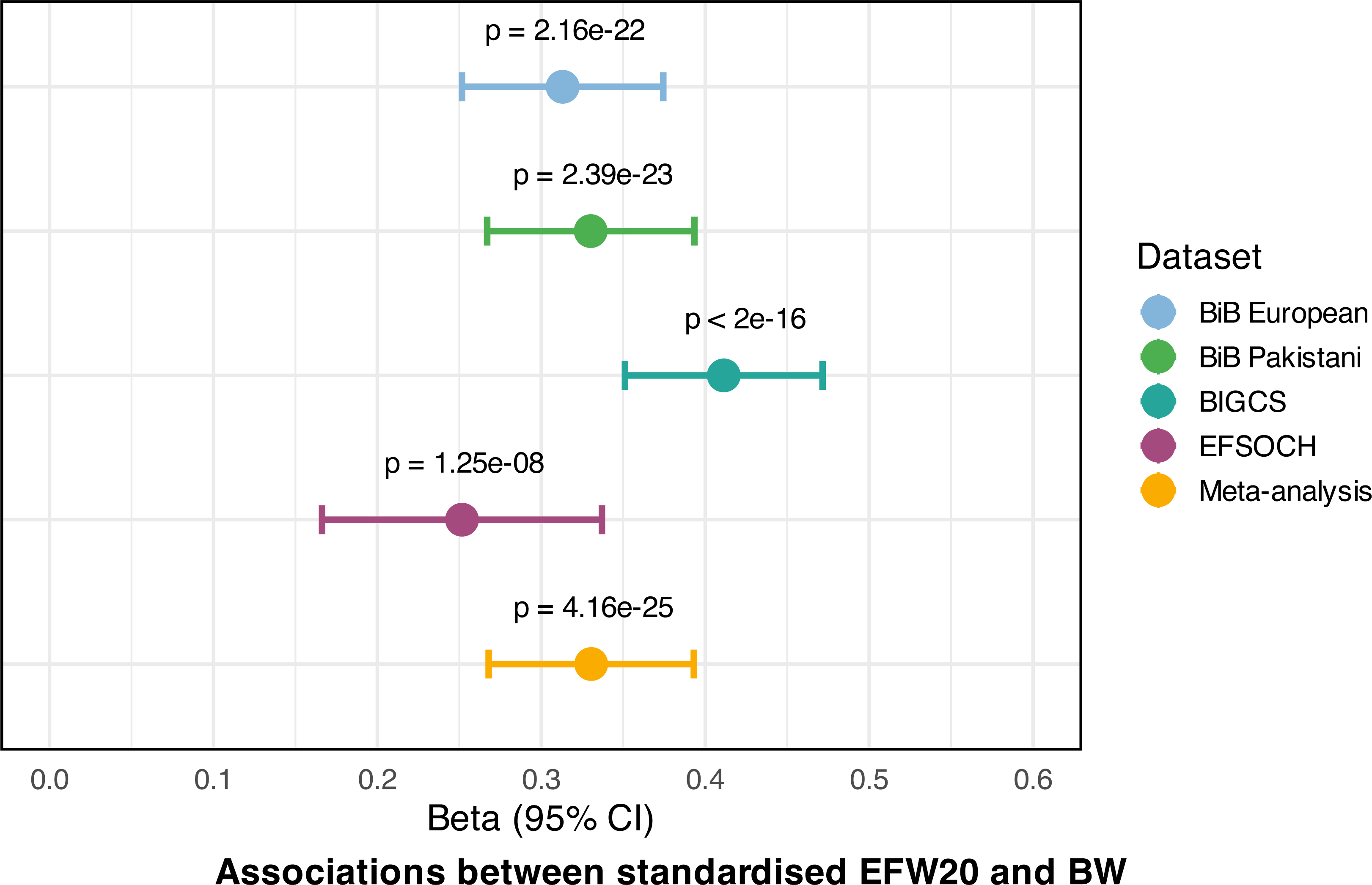
Association between EFW20 (standardized for gestational age at scan) and BW (standardized for gestational age at birth) with p-values for association displayed.

**Figure S3.**
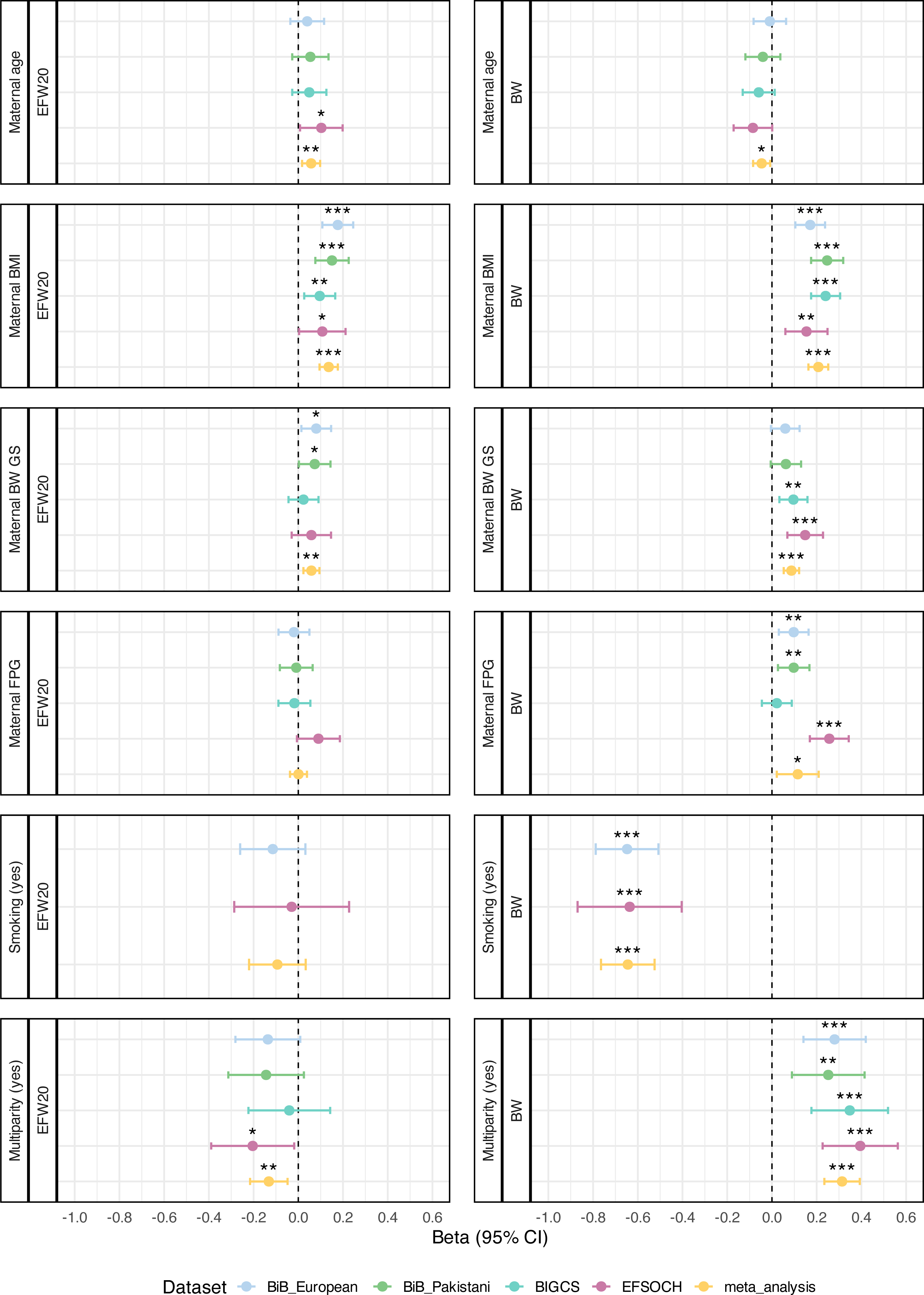
Sensitivity analysis of associations between maternal environmental factors and fetal growth outcomes at 20 weeks gestation and at term. Beta value represents standard deviation unit change in grams of estimated fetal weight at 20 weeks (EFW20) or birthweight (BW) per 1 unit change in the standardized or binary exposure variable. * = *p* <0.05, ** = *p* <0.01 and *** = *p* <0.001. Maternal smoking was not included in the BiB Pakistani and BIGCS maternal models due to low sample size of smokers.

