## Supporting Information Text 1 to 4 for "Genetic and maternal environmental contributions to estimated fetal weight at 20 weeks gestation compared with birthweight"

**Methods**

**Text 1: Genotyping, imputation and kinship analyses for cohorts.**

Born in Bradford (BiB) is a longitudinal multi-ethnic birth cohort study established in Bradford, UK, in 2007 to examine the impact of genetic, environmental, behavioural and social factors on child and maternal health and well-being in a deprived population.^1^ Data were originally collected from 12 453 women with 13 776 pregnancies, approximately half of whom were people of South Asian ethnicity (predominantly Pakistani), and half were non-South Asian ethnicity (predominantly White British)^1^. Genome-wide SNP genotyping and imputation were performed on a subset of individuals and have been described in detail previously.^2^ Following kinship analysis, up to third-degree relatives were excluded,^2^ totalling 4081 pregnancies in unrelated mothers with genotype data available for both mother and child.

The Exeter Family Study of Childhood Health (EFSOCH) is a prospective community-based study within central Exeter, UK, established in 2000 to investigate genetic and environmental influences on fetal growth and early development. Established inclusion criteria yielded a homogenous cohort of 1017 families (mother-father-child triads), comprised of non-diabetic mothers and fathers of self-reported UK Caucasian ethnicity.^3^ Genome-wide SNP genotyping and imputation have been described in detail previously.^4^ Kinship analysis was estimated using KING software.^5^ A total of 675 unrelated pregnancies had genotype data available for both mother and child.

The Born in Guangzhou Cohort Study (BIGCS) is a longitudinal birth cohort study based in the city of Guangzhou, southern China, launched in 2012 to explore how early-life traits impact developmental health.^6^ Data were originally collected from over 63 000 pregnancies from individuals of Chinese nationality. Genetic ancestry was assessed using self-reporting and cross-referenced with principal component (PC) analysis. Genotyping and imputation were performed using low-coverage whole-genome sequencing on a subset of participants, which have been described in detail previously.^7^ A total of 3851 pregnancies had genotype data available for both mother and child.

**Text 2: Hadlock’s Formula for estimated fetal weight.**

EFW = 10^(1.326 + 0.0107×HC + 0.0438×AC + 0.158×FL − 0.00326×AC×FL)

**Equation 1.** Hadlock’s formula.^8^ EFW: estimated fetal weight; HC: head circumference; AC: abdominal circumference; FL: femur length. Measurements of HC, AC and FL are all in centimetres.

**Text 3: Generation of birthweight genetic scores (BW GSs).**

We calculated fetal and maternal genetic scores for birthweight (BW GSs) by summing birthweight-raising alleles of 216 SNPs in each individual and weighting by the beta values that were reported with robust evidence of genome-wide significance in the largest published GWAS meta-analysis of birthweight^9^ and available in all the BiB, BIGCS, and EFSOCH study samples (see ST1). We generated the maternal and fetal-specific beta values using a weighted linear model (WLM)^10, 11^ that accounted for the correlation between maternal and fetal genotypes.^10^ We estimated the overlap between maternal and fetal genotypes in the birthweight GWAS summary statistics^9^ with LD score regression using LDSC v1.0.1^12^ and used the intercept in the WLM to produce fetal and maternal specific beta values for our SNPs of interest. These now adjusted-fetal and -maternal beta values became the 𝑤𝑖 used to generate the fetal and maternal genetic scores. Genetic scores were calculated using Equation 2, where 𝑤𝑖 is the beta value (fetal or maternal specific effect estimate) for SNP 𝑖, and 𝑔𝑖 is the dosage of the birthweight-raising allele at SNP 𝑖:

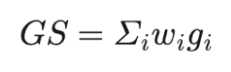

**Equation 2.** Genetic score calculation based on weight and dosage of effect alleles.

**Text 4: Sensitivity analyses to assess the influence on our results of gestational diabetes or the inclusion of ultrasound scan data later than 24 weeks.**

We conducted a sensitivity analysis with more stringent maternal glucose criteria, to control for women who may have had their glucose levels lowered after the point of the OGTT as they were identified as having gestational diabetes mellitus (GDM). As a result, mothers with glucose levels exceeding the threshold for a diagnosis of GDM (FPG of ≥5.6 mmol/L and 2-hour post-load OGTT levels of ≥7.8 mmol/L)^13^ were excluded. Women in the BiB and BIGCS cohorts with no 2-hr glucose data available were excluded. We performed an additional sensitivity analysis, repeating multivariable regression analyses for EFW20 in the sample of pregnancies with ultrasound scans restricted to 18-24 weeks in BIGCS. Gestational ages later than 24 weeks include the nutritionally and therefore placentally driven growth period,^14^ and this additional analysis was intended to draw a distinction between this later stage, captured by birthweight, and earlier fetal growth.
